# Prevalence and mortality associations of interstitial lung abnormalities in rheumatoid arthritis within a multicenter prospective cohort of smokers

**DOI:** 10.1101/2022.12.17.22283610

**Authors:** Gregory C McDermott, Keigo Hayashi, Kazuki Yoshida, Matthew Moll, Michael H Cho, Tracy J Doyle, Gregory L Kinney, Paul F Dellaripa, Rachel K Putman, Raul San Jose Estepar, Akinori Hata, Takuya Hino, Tomoyuki Hida, Masahiro Yanagawa, Mizuki Nishino, George Washko, Elizabeth Regan, Hiroto Hatabu, Gary M Hunninghake, Edwin K Silverman, Jeffrey A Sparks

**Author notes:** **Corresponding Author:** Jeffrey A. Sparks, MD, MMSc, Division of Rheumatology, Inflammation, and Immunity, Brigham and Women’s Hospital, 60 Fenwood Road, #6016U Boston, MA 02115, @jeffsparks. I, the Submitting Author has the right to grant and does grant on behalf of all authors of the Work (as defined in the below author licence), an exclusive licence and/or a non-exclusive licence for contributions from authors who are: i) UK Crown employees; ii) where BMJ has agreed a CC-BY licence shall apply, and/or iii) in accordance with the terms applicable for US Federal Government officers or employees acting as part of their official duties; on a worldwide, perpetual, irrevocable, royalty-free basis to BMJ Publishing Group Ltd (“BMJ”) its licensees and where the relevant Journal is co-owned by BMJ to the co-owners of the Journal, to publish the Work in Annals of the Rheumatic Diseases and any other BMJ products and to exploit all rights, as set out in our <u>licence</u>. The Submitting Author accepts and understands that any supply made under these terms is made by BMJ to the Submitting Author unless you are acting as an employee on behalf of your employer or a postgraduate student of an affiliated institution which is paying any applicable article publishing charge (“APC”) for Open Access articles. Where the Submitting Author wishes to make the Work available on an Open Access basis (and intends to pay the relevant APC), the terms of reuse of such Open Access shall be governed by a Creative Commons licence – details of these licences and which <u>Creative Commons</u> licence will apply to this Work are set out in our licence referred to above.

## Abstract

**Objectives:** Investigate the prevalence and mortality impact of interstitial lung abnormalities (ILA) in rheumatoid arthritis (RA) and non-RA comparators.

**Methods:** We analyzed associations between ILA, RA, and mortality in COPDGene, a multicenter prospective cohort study of current or former smokers, excluding known interstitial lung disease (ILD) or bronchiectasis. All participants had research chest high-resolution computed tomography (HRCT) reviewed by a sequential reading method to classify ILA as present, indeterminate, or absent as well as fibrotic or nonfibrotic ILA subtype. RA cases were identified by self-report RA and DMARD use; non-RA comparators had neither an RA diagnosis nor used DMARDs. We examined the association and mortality risk of RA and ILA using multivariable logistic regression and Cox regression.

**Results:** We identified 83 RA cases and 8725 non-RA comparators with HRCT performed for research purposes. ILA prevalence was 16.9% in RA cases and 5.0% in non-RA comparators. After adjusting for potential confounders including genetics, smoking, and other lifestyle factors, ILA were more common among those with RA compared to non-RA (OR 4.76 95%CI 2.54 to 8.92). RA with ILA or indeterminate for ILA was associated with higher mortality compared to non-RA without ILA (HR 3.16, 95%CI 2.11 to 4.74) and RA cases without ILA (HR 3.02, 95%CI 1.36 to 6.75).

**Conclusions:** RA was associated with ILA and this persisted after adjustment for smoking and genetic/lifestyle risk factors. RA with ILA in chronic heavy smokers had 3-fold increased mortality, emphasizing the importance of further screening and treatment strategies for subclinical ILD in RA.

**KEY MESSAGES:** *What is already known on this topic:*

- Up to a third of patients with rheumatoid arthritis (RA) may have evidence of subclinical interstitial lung abnormalities on computed tomography (CT) scans of the chest.
- Cigarette smoking and the *MUC5B* promoter variant are known risk factors for RA-associated interstitial lung disease.

*What this study adds:*

- We found that 17% of RA patients had subclinical interstitial lung abnormalities. RA had 4-fold higher odds of interstitial lung abnormalities than non-RA comparators, adjusted for smoking, the *MUC5B* promoter variant, and other factors.
- Participants with RA and no interstitial lung abnormalities were not at increased mortality risk while those with interstitial lung abnormalities or indeterminate for ILA had a three-fold increased risk of mortality compared to RA and non-RA patients without interstitial lung abnormalities.

*How this study might affect research, practice or policy:*

- The presence of subclinical interstitial lung abnormalities confers significant mortality risk in RA and emphasizes the need to establish the clinical utility of screening, prevention, and treatment strategies targeting subclinical lung disease.

## INTRODUCTION

Rheumatoid arthritis-associated interstitial lung disease (RA-ILD) is a severe extra-articular disease manifestation and a major driver of morbidity and mortality among RA patients.^1–7^ Although 5-10% of RA patients may develop clinically-apparent RA-ILD, research screening studies have demonstrated that up to a third of RA patients may have subclinical ILD based on evaluations of high-resolution computed tomography (HRCT) scans.^1,8–12^ RA shares several risk factors with idiopathic pulmonary fibrosis, the stereotypical fibrotic interstitial lung disease, including cigarette smoking, aging, and male sex.^8,13^ Thus, it remains unknown whether the prevalence of lung abnormalities in RA simply reflects these overlapping risk factors or is explained by RA-related pulmonary inflammation, RA treatment effects, or other mechanisms. Additionally, the prognostic implications of subclinical pulmonary abnormalities in RA remain unclear, as only a minority of RA patients with pulmonary abnormalities will develop clinically apparent lung disease.^14^

Although there has been limited previous research of the implications of subclinical lung disease in RA, several large cohort studies have investigated subclinical pulmonary abnormalities in the general population.^15–17^ These studies have largely focused on interstitial lung abnormalities (ILA), a term that refers to specific radiologic patterns of increased lung density noted on HRCT chest imaging.^18,19^ ILA share several characteristics and risk factors with idiopathic pulmonary fibrosis and other forms of ILD, and may represent a preclinical form of these conditions in some patients. ILA can occur in up to 7% of the general population^17^ and have been associated with increased mortality in multiple cohorts.^15,16^ There have also been some investigations of ILA in the RA population. One prior study of RA patients without known ILD noted that 15% had ILA on screening HRCT.^20^ Other studies investigating RA patients with clinically-performed CT scans have noted a prevalence of subclinical interstitial lung disease as high as 44-45%.^14,21^ Other studies, which did not specifically use an ILA definition, found HRCT evidence of subclinical interstitial lung disease in 16.9-33% of RA patients.^9,10,22,23^ However, these studies did not specifically include non-RA controls and address known ILA risk factors, including age, smoking status, and the promoter variant of the *MUC5B* gene.

We used data from a large prospective cohort of current and former smokers to determine the prevalence of ILA and investigate the long-term mortality risk of ILA among patients with and without RA. We hypothesized that RA would be associated with ILA, even after adjustment for smoking and other risk factors. We further hypothesized that the combination of RA and ILA would confer significantly higher mortality risk than either condition alone.

## METHODS

### Study population and design

We analyzed COPDGene, a multicenter prospective cohort of current or former smokers that has been described in detail elsewhere.^24–26^ Briefly, the study enrolled non-Hispanic White or Black smokers with at least 10 pack-years of smoking history, aged 45-80 years, at 21 clinical centers in the United States between 2007-2011. The study enrolled smokers with and without baseline obstructive lung disease and reached its goal to include one-third Black participants. Baseline screening included health questionnaires, HRCT chest imaging, spirometry, and genotyping. Participants with known respiratory diseases besides asthma, and those with significant ILD or bronchiectasis on chest CT, were ineligible. We performed a cross-sectional study investigating the association of RA compared to non-RA with ILA. We then performed a cohort study examining the associations of RA and ILA with mortality risk. This substudy was approved by the Mass General Brigham Institutional Review Board (protocol 2020P000558).

### RA cases and non-RA comparators

Within COPDGene, we identified participants with RA based on a combination of self-reported physician-diagnosed RA and use of at least one disease-modifying rheumatic medication (DMARD) at baseline. A previous study noted low validity of self-report RA status alone but reported a positive predictive value (PPV) of 88% using a similar case definition that combined self-report status with DMARD use.^27^ We included medications approved for RA by the US Food and Drug Administration at the time COPDGene baseline visits were conducted and other DMARDs previously validated for identification of RA patients in cohort studies (**Supplemental Table S1**).^28^ We defined non-RA comparators as participants who did not report a history of RA and were not using any DMARDs at baseline. Participants who reported RA history but did not report DMARD use and those on one or more DMARDs without reporting a history of RA were excluded as comparators.

### Interstitial lung abnormalities

The presence of ILA on the research chest HRCT was identified using a previously described sequential reader review of HRCT images by up to three expert readers (two chest radiologists and one pulmonologist) to classify ILA as present, indeterminate, or absent.^24,29^ ILA was defined as nondependent subpleural changes affecting >5% of any lobar area. These included nondependent ground-glass and reticular opacities, diffuse centrilobular nodules, nonemphysematous cysts, honeycombing, and traction bronchiectasis. ILA were further subcategorized into fibrotic and nonfibrotic subtypes, consistent with recent guidelines that recognized a higher rate of progression and poorer prognosis among those with fibrotic ILA.^18^ Scans with focal or unilateral changes, or patchy ground-glass opacities affecting <5% of a lobar region were considered indeterminate for ILA.

### Chronic Obstructive Pulmonary Disease (COPD)

We used each participant’s post-bronchodilator forced expiratory volume in 1 second (FEV_1_) and forced vital capacity (FVC) to categorize them into severity grades according to the Global Initiative for Chronic Lung Disease 2007 classification system.

### Mortality

Death outcomes in COPDGene were assessed through longitudinal study follow-up by biannual phone calls and periodic searches of the social security death index. The mortality dataset was last updated in August 2022.

### Covariates

We obtained age at COPDGene baseline visit, sex, and self-reported race. Body mass index (BMI) was obtained from the baseline examination. Smoking status (current, former) and pack-years of smoking were obtained from baseline respiratory health questionnaire. As the *MUC5B* promoter variant rs35705950 has been associated with RA in some cohorts^30^ and is the strongest known genetic risk factor for ILA and RA-ILD^17,31^, this genetic risk factor was included as a covariate. Patients were genotyped using the TaqMan real time polymerase chain reaction assay (Thermo Fisher Scientific, Waltham, MA). To minimize the possibility of confounding by ancestry in analyses that included this genetic marker, we used genotyping data to determine principal components of ancestry and included the first six principal components of ancestry as additional covariates.

### Statistical analysis

We compared baseline characteristics between RA cases and non-RA comparators using frequencies, proportions, and means with standard deviations. We examined the association between RA case vs non-RA comparator status with ILA, indeterminate for ILA, and the fibrotic ILA subtype using unadjusted and multivariable logistic regression adjusted for age, sex, smoking status, pack-years of smoking, and BMI. We performed an additional model that also adjusted for genetic ancestry and the *MUC5B* rs35705950 genotype, treating the number of risk alleles for each patient as an ordinal variable. Participants with missing *MUC5B* promoter variant genotype (n=5 RA cases and n=259 non-RA comparators) were excluded from that analysis. We assessed for a possible association between RA and COPD, as defined by GOLD class 2-4, using multivariable logistic regression adjusted for age, sex, smoking status, and pack-years of smoking. There were 51 non-RA comparators who were missing spirometry data and excluded from these analyses. There were no other missing data.

For the mortality analyses, we constructed Kaplan-Meier curves with cumulative incidence of mortality based on RA and ILA status. We also performed Cox regression models to estimate hazard ratios for mortality based on RA vs. non-RA status as well as a cross-classified variable based on ILA presence/absence and RA/non-RA status. We performed unadjusted models as well as multivariable models adjusted for age, sex, smoking status, pack-years, and BMI. We also performed these analyses limited to RA cases. In secondary analyses, we repeated the Cox proportional hazards models after excluding indeterminate for ILA and another analysis after excluding ILA.

We assessed the proportional hazards assumptions in all analyses using Schoenfeld residuals. There were no significant violations of the proportional hazards assumptions except for the age and BMI covariates, which showed interactions with follow up time and the mortality outcome. In models that included interaction terms of these variables with follow up time, the point estimates of the main exposures were unchanged so we reported results without the interaction terms. Two-sided p values of <0.05 were considered statistically significant. All analyses were performed using SAS v9.4 (Cary, NC). Patients and the public were not involved in the design or implementation of this substudy.

## RESULTS

### Study sample

Out of 10,371 COPDGene participants, we identified 83 RA cases and 8,725 non-RA comparators scored for ILA (**Figure 1**). RA cases self-reported a history of RA and were using at least one DMARD at baseline. Details of specific DMARD use are detailed in **Supplemental Table S1**. A total of 656 participants who reported a history of RA but were not using DMARDs and 45 participants who reported DMARD use but no RA history were excluded. We also excluded 2 participants with RA and 611 non-RA comparators who were not scored for ILA.

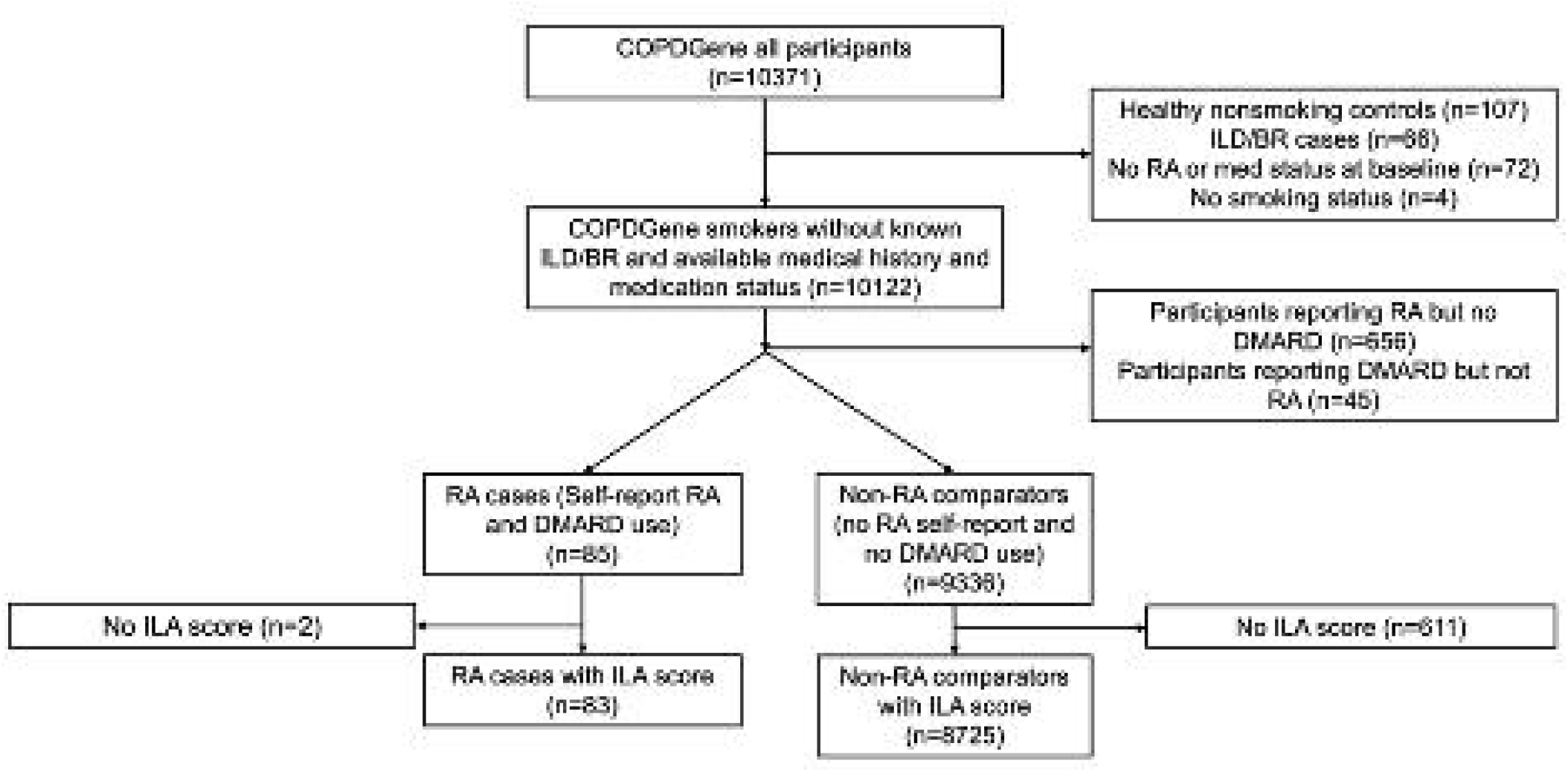

### Baseline characteristics

Baseline characteristics of RA and non-RA participants are shown in **Table 1**. Participants with RA were older at enrollment (63.8 vs 59.3 years) and more predominantly female (65.1 vs. 45.9%) and White race (79.5 vs 67.5%). The RA cases had a higher proportion of former smokers (66.3 vs 46.8%) and had higher mean BMI (30.0 vs 28.7 kg/m^2^).

**Table 1:** Select demographics, lifestyle factors, pulmonary disease, and genetics of RA patients and non-RA comparators at baseline (n=8808).

|  | **RA cases**<br>**(n=83)** | **Non-RA comparators**<br>**(n=8725)** |
| --- | --- | --- |
| **Demographics** |  |  |
| Age at enrollment (years, mean, SD) | 63.8 (8.7) | 59.3 (9.0) |
| Female (n, %) | 54 (65.1%) | 4002 (45.9%) |
| White race (n, %) | 66 (79.5%) | 5890 (67.5%) |
| Black race (n, %) | 17 (20.5%) | 2835 (32.5%) |
| **Lifestyle** |  |  |
| Current smoker (n, %) | 28 (33.7%) | 4645 (53.2%) |
| Past smoker (n, %) | 55 (66.3%) | 4080 (46.8%) |
| Pack-years (mean, SD) | 42.2 (17.9) | 44.0 (24.6) |
| BMI (kg/m<sup>2</sup>, mean, SD) | 30.0 (7.8) | 28.7 (6.2) |
| **MUC5B promoter variant (rs35705950) genotype** |  |  |
| GG | 63/78 (80.8%) | 7266/8466 (85.8%) |
| GT | 15/78 (19.2%) | 1136/8466 (13.4%) |
| TT | 0/78 (0.0%) | 64/8466 (0.8%) |
| **Chronic obstructive pulmonary disease\*** |  |  |
| GOLD class 0 (no COPD) | 32 (38.6%) | 3790/8674 (43.7%) |
| PRISm | 13 (15.7%) | 1063/8674 (12.3%) |
| GOLD class 1 | 3 (3.6%) | 697/8674 (8.0%) |
| GOLD class 2 | 17 (20.5%) | 1627/8674 (18.8%) |
| GOLD class 3 | 13 (15.7%) | 986/8674 (11.4%) |
| GOLD class 4 | 5 (6.0%) | 511/8674 (5.9%) |

BMI = body mass index; COPD = chronic obstructive pulmonary disease; GOLD = Global Initiative for Chronic Obstructive Lung Disease; ILA = interstitial lung abnormalities; PRISm = Preserved Ratio Impaired Spirometry (subset of GOLD class 0); RA = rheumatoid arthritis; SD = standard deviation
\* = based on GOLD 2007 classification using postbronchodilator FEV<sub>1</sub>/FVC only

### COPD associations

The proportion of RA cases and non-RA comparators in each GOLD class was similar (**Table 1**). There were no statistically significant associations between RA and COPD. Odds ratios (OR) were 1.21 (95%CI 0.78 to 1.88) in the unadjusted analysis and 0.93 (95%CI 0.58 to 1.50) in the analysis adjusted for age, sex, smoking status, and pack-years (**Supplemental Table S2**).

### ILA prevalence and associations

A higher proportion of RA cases had ILA (16.9%) compared to the non-RA comparators (5.0%, **Figure 2**). Additionally, 8.4% of the RA cases had fibrotic ILA compared to 1.1% of the non-RA comparators. There was also a higher proportion of participants who were indeterminate for ILA among the RA cases (41.0%) compared to those without RA (35.8%).

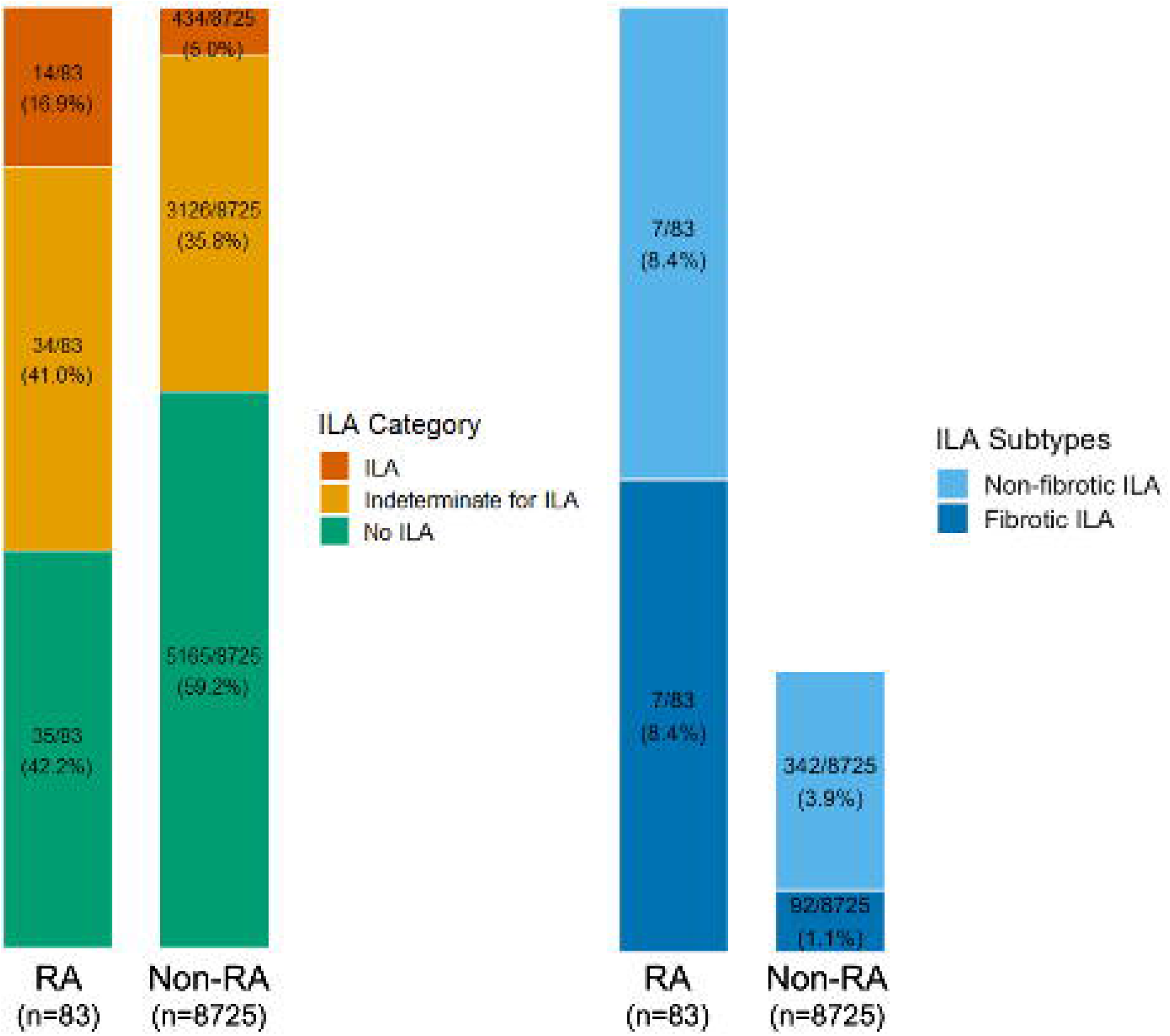

In the multivariable logistic regression model adjusted for age, sex, smoking status, pack-years, and BMI, RA cases had an OR of 4.15 (95%CI 2.17 to 7.97) for ILA compared to those without RA (**Table 2**). For RA cases, the OR for fibrotic ILA was 9.26 (95%CI 3.82 to 22.44). Results were similar after adjustment for the number of risk alleles for the *MUC5B* promoter variant (rs35705950), as shown in **Table 2**.

**Table 2:** Cross-sectional association of RA and radiologic interstitial lung abnormalities (ILA) on HRCT at baseline (n=8808).

|  | **Unadjusted OR**<br>**(95%CI)** | **Model 1\* OR**<br>**(95%CI)** | **Model 2† OR**<br>**(95%CI)** |
| --- | --- | --- | --- |
| *Outcome: ILA or indeterminate for ILA vs. no ILA* |  |  |  |
| Non-RA comparators | 1.0 (Ref) | 1.0 (Ref) | 1.0 (Ref) |
| RA cases | 1.99 (1.28 to 3.08) | 1.87 (1.20 to 2.92) | 1.84 (1.16 to 2.92) |
| *Outcome: ILA vs. no ILA* |  |  |  |
| Non-RA comparators | 1.0 (Ref) | 1.0 (Ref) | 1.0 (Ref) |
| RA cases | 4.76 (2.54 to 8.92) | 4.15 (2.17 to 7.97) | 3.90 (1.95 to 7.80) |
| *Outcome: Fibrotic ILA vs. no ILA* |  |  |  |
| Non-RA comparators | 1.0 (Ref) | 1.0 (Ref) | 1.0 (Ref) |
| RA cases | 11.23 (4.86 to 25.94) | 8.41 (3.49 to 20.25) | 8.18 (3.17 to 21.08) |

\* adjusted for age, sex, smoking status (current/past), pack-years, and body mass index.
† additionally adjusted for *MUC5B* rs35705950 genotype (copy number of T risk alleles; missing excluded) and the first six principal components of ancestry
HRCT = high resolution computed tomography; ILA = interstitial lung abnormalities; RA = rheumatoid arthritis

### RA, ILA, and mortality risk

In the mortality analysis, the median follow-up time was 10.5 years (IQR 5.8, 12.4) for RA cases and 10.7 years (IQR 5.4, 12.4) for non-RA comparators. There were 33 deaths among the RA cases and 2,225 among non-RA comparators during 740.9 and 76,594.5 person years, respectively (**Supplemental Table S3**). The 10-year mortality was 50.0% (95%CI 36.4 to 64.6%) in participants with RA and ILA or indeterminate for ILA compared to 22.0% (95%CI 7.5 to 36.5%) for RA cases without ILA and 21.2% (95%CI 20.6 to 21.9%) for non-RA participants without ILA.

Kaplan-Meier cumulative mortality curves stratified by RA and ILA status are shown in **Figure 3, Figure 4A and 4B**. In the Cox regression models adjusted for age, sex, smoking status, pack-years, and BMI, RA had a HR for mortality of 1.52 (95%CI 1.07 to 2.14) as shown in **Figure 3**. After stratifying by ILA status, the HR for mortality was 0.89 (95%CI 0.24 to 3.27) for the RA participants with ILA and 4.21 (95%CI 1.03 to 9.20) for the RA participants with indeterminate for ILA findings, compared to non-RA participants without ILA (**Figure 4**). When we combined RA with ILA or indeterminate for ILA and compared to participants without ILA, the group with ILA/indeterminate for ILA had a hazard ratio (HR) of 2.64 (95%CI 1.76 to 3.97) for death compared to participants without RA or ILA (**Figure 5**). When we limited our analysis to RA cases, RA cases with ILA or indeterminate for ILA findings had HR of 2.86 (95%CI 1.33 to 6.16) compared to RA cases without ILA. There were no significant differences in HR for mortality comparing RA and non-RA participants without ILA (HR 0.92 95%CI 0.48 to 1.78).

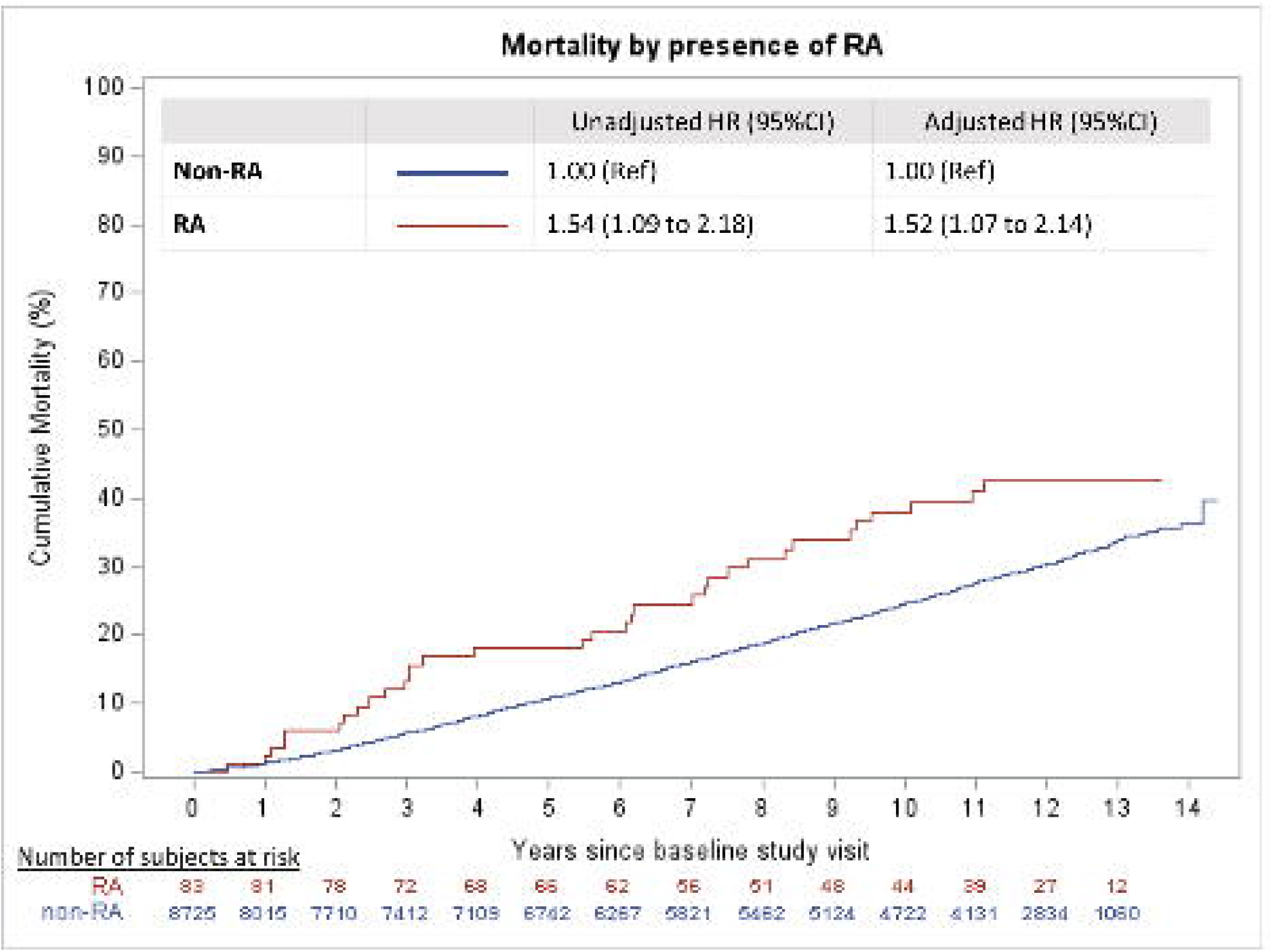

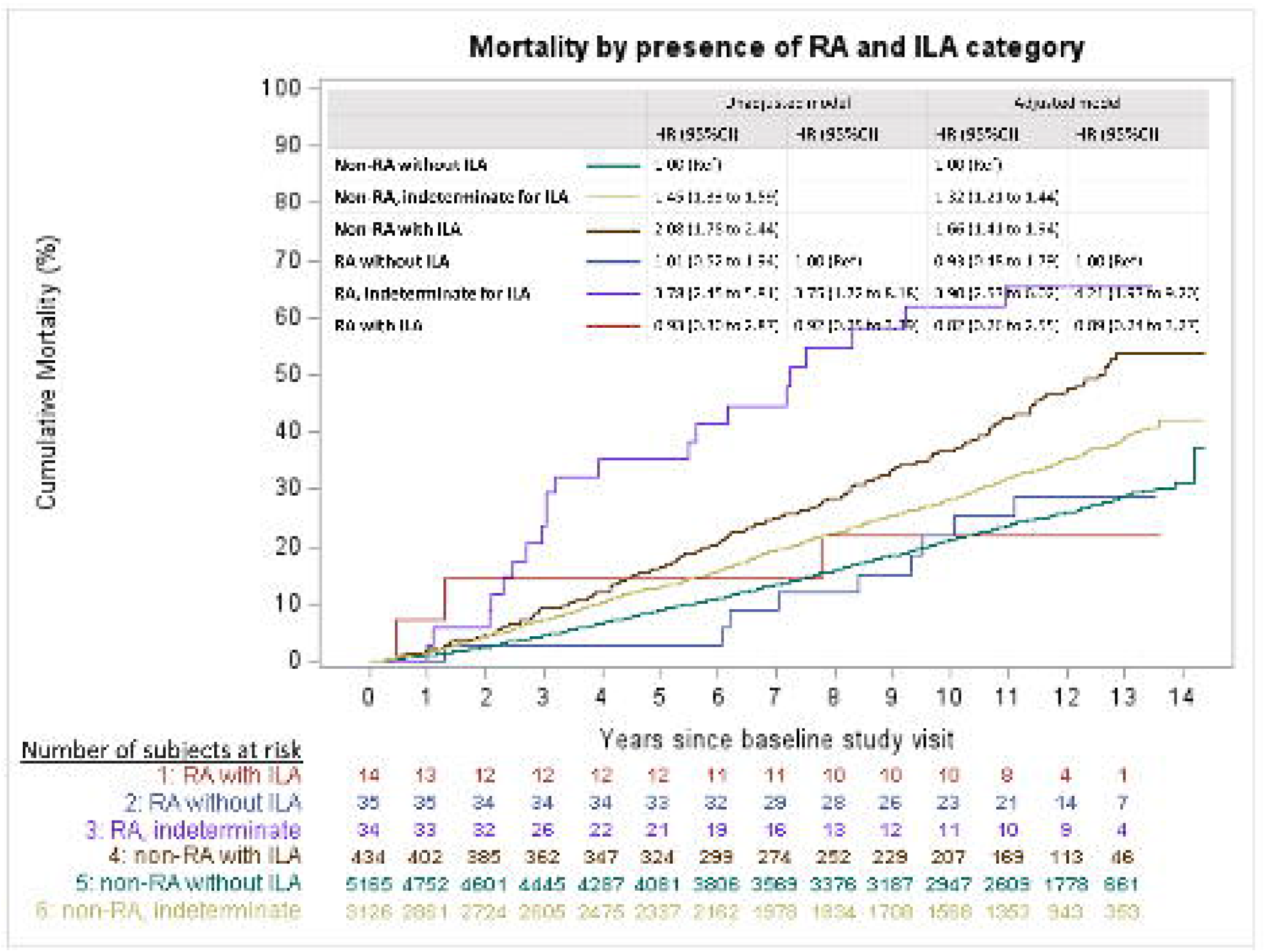

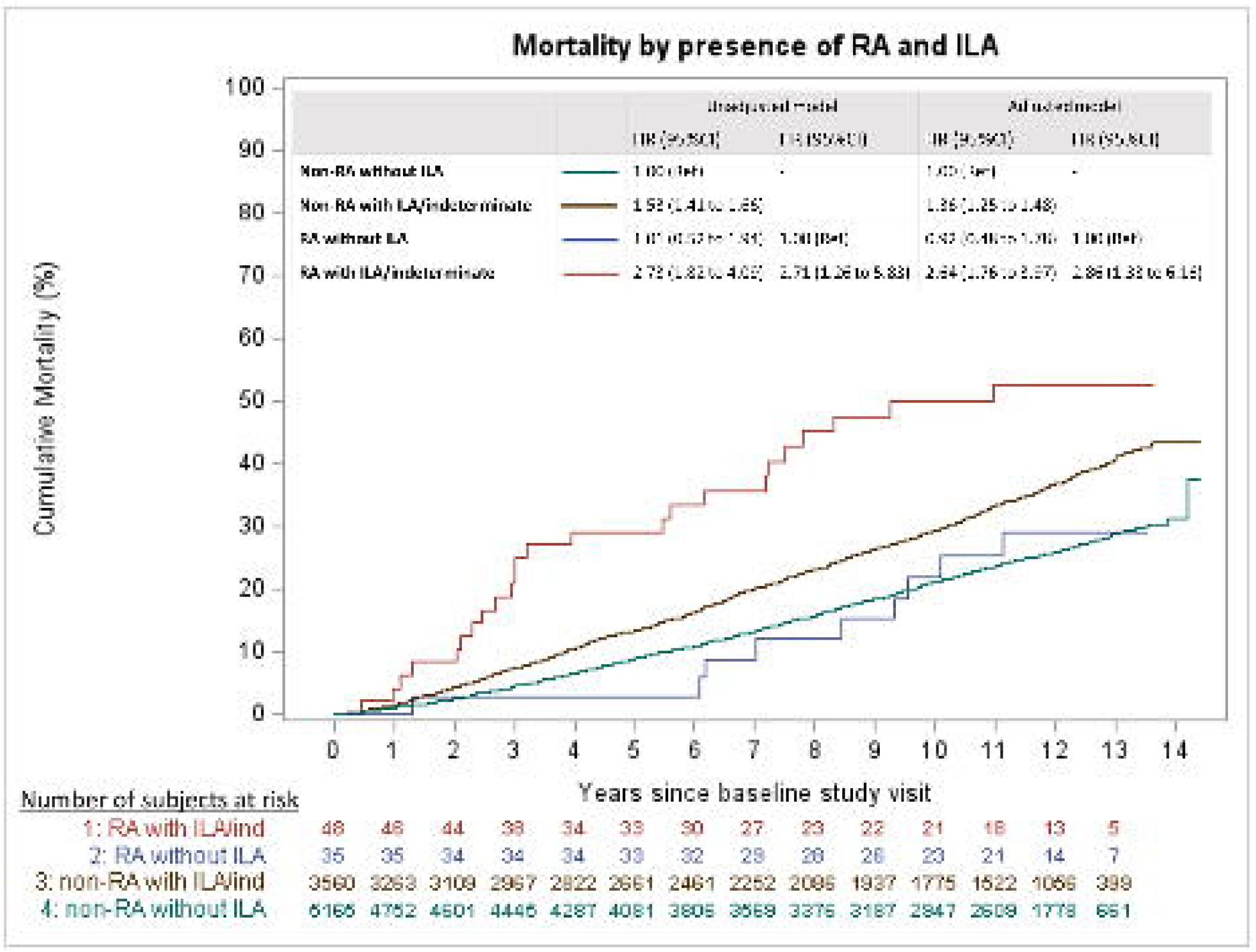

## DISCUSSION

In this large comparative study, RA was strongly associated with ILA, even after adjustment for cigarette smoking, the *MUC5B* promoter variant, and other risk factors. The prevalence of ILA in RA was 17% compared to 5% in non-RA in this cohort composed of smokers without clinical evidence of ILD or bronchiectasis. Furthermore, we found that RA with ILA or indeterminate for ILA had 3-fold increased mortality compared to those without ILA. These findings suggest that interstitial lung abnormalities have important prognostic significance in RA patients and emphasize the urgent need for additional research investigating the screening, monitoring, and early treatment strategies for subclinical ILD in RA, especially in high-risk patients with significant smoking history.

Our study adds to the existing literature investigating subclinical lung disease in RA. One study of 177 RA patients in the ESCAPE RA cohort found that 32% had at least one ILD feature on cardiac CT scan.^32^ However, this study did not include non-RA comparators. An additional investigation compared ESCAPE RA patients to non-RA controls in the Multi-Ethnic Study of Atherosclerosis (MESA) and found evidence of an association between RA and increased high attenuation areas in the lung parenchyma captured on cardiac CT scans.^33^ The MESA study also noted an association between high attenuation areas and RA-related antibodies in a separate study.^34^ However, these studies did not investigate ILA, use dedicated full lung HRCT imaging, or specifically exclude patients with known ILD. Several additional prospective studies have investigated the prevalence of subclinical lung disease in RA patients using lung HRCT.

One screening study in the BRASS registry performed research HRCT scans on RA patients without known ILD diagnosis and found evidence of ILA in 15%.^35^ Another recent investigation performed in the French ESPOIR and TRANSLATE2 cohorts obtained HRCT on RA patients without ILD history, respiratory symptoms, or signs of pulmonary disease on examination.^22^ This study noted subclinical ILD in 16.9% and 19.0% of the subjects. Similar investigations performed in small cohorts of RA patients without known lung disease at the National Institutes of Health in Maryland and Cedars Sinai in California noted subclinical ILD on HRCT in 33% and 39%, respectively.^36,37^ Perhaps not surprisingly, the prevalence of subclinical ILD may be higher on HRCT performed for clinical purposes. A separate study performed in the BRASS cohort used a sequential reader method to assess for evidence of ILA on clinically performed CT scans of RA patients. This investigation noted evidence of ILA changes in 45% of RA patients, including 40% of patients without a known history of pulmonary fibrosis.^21^ These studies did not include non-RA comparators or account for potential risk factors like smoking and the *MUC5B* promoter variant.

Although the prevalence of subclinical lung abnormalities has been well established in RA patients, less is known about the progression of these abnormalities and their impact on mortality. One study from Cedars-Sinai noted progression of interstitial abnormalities in 86% of patients after one year.^37^ A retrospective investigation performed in Brazil noted progression of ILA in 29% of patients that had clinical testing performed.^14^ However, neither study included non-RA comparators. Furthermore, although risk factors for progression of ILA have been identified in the general population, including the fibrotic subtype of ILA, risk factors for ILA progression in RA remain largely unknown. One study did not find any impact of the *MUC5B* promoter variant on ILD progression in RA patients.^38^ Furthermore, although multiple studies have demonstrated increased mortality associated with clinically-apparent RA-ILD, none have investigated the impact of subclinical RA-ILD on mortality. Our study noted a strong association between subclinical ILA and mortality among RA patients. Interestingly, the mortality was highest in the RA group that was indeterminate for ILA and we did not see a significant association between ILA and mortality in the RA subgroup. This may be due to the small sample size of the RA with ILA subgroup or perhaps related to some other feature of the “indeterminate for ILA” designation that leads to higher mortality. Additional larger studies investigating this issue are needed.

Our study offers epidemiologic evidence of an important link between RA and ILA and found a particularly strong association with fibrotic ILA. The mucosal origins hypothesis for RA pathogenesis posits that damage to the airway and respiratory mucosa leads to aberrant inflammation, dysbiosis, citrullination of peptides, and the generation of RA-related autoantibodies.^39^ Evidence for this hypothesis derives from several studies that have noted evidence of pulmonary inflammation and RA-related autoantibodies in sputum samples prior to the onset of joint inflammation.^34,40^ Despite recognition of the important role of the lung as a mucosal site of autoantibody production, we have limited understanding of why only a minority of RA patients with ILA will progress to clinically-apparent RA-ILD. These mechanisms may relate to underlying genetic risk factors like the *MUC5B* promoter variant or involve ongoing injury to the pulmonary parenchyma through the direct effects of inflammation, medications, or recurrent respiratory infections, smoking, or other inhalants as a trigger in the setting of immunosuppression.^41,42^ Further studies are needed to characterize risk factors for progression and determine pathways that can be targeted by medications that arrest progression of lung disease.

Our study has several strengths. This investigation was performed using a large, prospective, longitudinal cohort that collected extensive details on smoking history and respiratory risk factors. We specifically investigated presence of ILA in RA compared to non-RA and carefully adjusted for current smoking status and pack-years. Furthermore, we determined ILA status using a validated sequential reading method with expert reviewers who were blinded to the exposures of interest of this study. Lastly, there was a long duration of follow-up to examine mortality risk, and deaths were systematically captured by a combination of longitudinal follow-up and queries of the social security death index.

Our study also has several limitations to consider. First, we relied on a combination of self-report RA status and use of DMARD medications to identify participants with and without RA. However, other research studies have demonstrated a positive predictive value of 88% for RA using similar methods.^27,28^ Second, because COPDGene was not investigating RA, we have limited information on RA-related covariates. Thus, we were unable to examine how medication use, disease activity, serostatus, bone erosions, or RA duration may impact ILA risk in RA.^43^ However, we were able to account for important lifestyle, demographic, and genetic risk factors that are known risk factors for RA-ILD and may influence mortality.^31,44^ Third, since the COPDGene study was limited to White and Black smokers, our results may not be generalizable to other populations, including nonsmokers. However, smoking remains an important risk factor for RA development and smokers represent a high risk group that would be a likely target of future screening strategies.^6,12,37,44^ Fourth, despite the large sample size of the COPDGene cohort, the RA group was relatively small. While we were able to detect associations of RA with specific ILA subtypes such as fibrotic ILA, the mortality investigation was relatively limited due to the small number of deaths in this subgroup. Similarly, due to the size of our RA cohort, we were only able to study all-cause mortality, rather than cause-specific mortality.^12^ Larger studies with longer follow-up are needed to determine progression of specific ILA subtypes among RA patients.

In conclusion, we found that RA was strongly associated with ILA in this large comparative study. The association between RA and ILA persisted after adjustment for smoking intensity, smoking duration, and the *MUC5B* promoter variant. RA patients with ILA or indeterminate for ILA had 3-fold increased mortality compared to those without ILA. These findings emphasize the significance of ILA among RA patients and the need for further research to establish the clinical utility of screening, monitoring, and early treatment strategies for subclinical ILD.

## Data Availability

Data are available upon reasonable request and with appropriate institutional review board approval.

## Acknowledgements

We would like to acknowledge the COPDGene investigators, staff, and participants for their valuable contributions to this study.

## Disclaimer

The funders had no role in the decision to publish or in the preparation of this article. The content is solely the responsibility of the authors and does not necessarily represent the official views of Harvard University, its affiliated academic healthcare centres or the National Institutes of Health.

## Contributors

GCM and JAS had access to the study data, developed the figures and tables, and vouch for the data and analyses. KH performed the statistical analyses and contributed to data quality control, data analysis, and interpretation of the data. GCM, KH, MM, MHC, TJD, PFD, RKP, RS, GW, ER, HH, GMH, ES, and JAS contributed to data collection, data analysis, and interpretation of the data. JAS directed the work, designed the data collection methods, contributed to data collection, data analysis, and interpretation of the data and had final responsibility for the decision to submit for publication. All authors contributed intellectual content during the draft and revision of the work and approved the final version to be published.

## Patient and public involvement

Patients and the public were not involved in the design, conduct, reporting, or dissemination plans of this research.

## Patient consent for publication

Not required.

## Ethics approval

The study was approved by the Mass General Brigham Institutional Review Board, protocol number 2020P000558.

