## Supplemental Table S1 for "Prevalence and mortality associations of interstitial lung abnormalities in rheumatoid arthritis within a multicenter prospective cohort of smokers"

**Supplemental Table S1: Frequency of specific DMARD and glucocorticoid use among RA cases at COPDGene baseline (n=83)**

| **Medication** | **n (%)** |
| --- | --- |
| Methotrexate | 45 (54.2%) |
| TNF inhibitor | 27 (32.5%) |
| Etanercept | 15 (18.1%) |
| Infliximab | 9 (10.8%) |
| Adalimumab | 3 (3.6%) |
| Hydroxychloroquine | 21 (25.3%) |
| Sulfasalazine | 10 (12.0%) |
| Leflunomide | 5 (6.0%) |
| Azathioprine | 5 (6.0%) |
| Mycophenolate/mycophenolic acid | 1 (1.2%) |
| Abatacept | 1 (1.2%) |
| Glucocorticoids* | 23 (27.4%) |

Not mutually exclusive, some participants were on >1 DMARD. No participants reported minocycline, certolizumab pegol, golimumab, rituximab, tocilizumab, or anakinra use.

* Note: glucocorticoids were not considered a DMARD for the purposes of identifying RA cases and non-RA comparators

**Supplemental Table S2: Association of RA with COPD (GOLD class 2-4)**

|  | **Unadjusted OR (95% CI)** | **Multivariable Adjusted* OR (95% CI)** |
| --- | --- | --- |
| *Outcome: GOLD Class 2-4, GOLD Class 1 excluded* | | |
| Non-RA comparators | 1.0 (Ref) | 1.0 (Ref) |
| RA cases | 1.21 (0.78 to 1.88) | 0.93 (0.58 to 1.50) |
| *Outcome: GOLD Class 2-4, Gold Class 1 included* | | |
| Non-RA comparators | 1.0 (Ref) | 1.0 (Ref) |
| RA cases | 1.30 (0.84 to 2.01) | 1.02 (0.65 to 1.62) |

***** adjusted for age, sex, smoking status (current/former), pack-years

**Supplemental Table S3 Number of deaths, subjects, and person time stratified by RA/comparator and interstitial lung abnormality (ILA) status in COPDGene (n=8808)**

|  | **# of deaths** | **# of subjects** | **Total person-years** |
| --- | --- | --- | --- |
| Non-RA without ILA | 1129 | 5165 | 46584.5 |
| Non-RA, indeterminate for ILA | 917 | 3126 | 26387.4 |
| Non-RA with definite ILA | 179 | 434 | 3622.6 |
| RA without ILA | 9 | 35 | 364.7 |
| RA, indeterminate for ILA | 21 | 34 | 243.0 |
| RA with definite ILA | 3 | 14 | 133.2 |

**Supplemental Figure S1 Cumulative mortality and hazard ratios stratified by RA/comparator and interstitial lung abnormalities (ILA) status in COPDGene, indeterminate for ILA excluded (n=5823)**

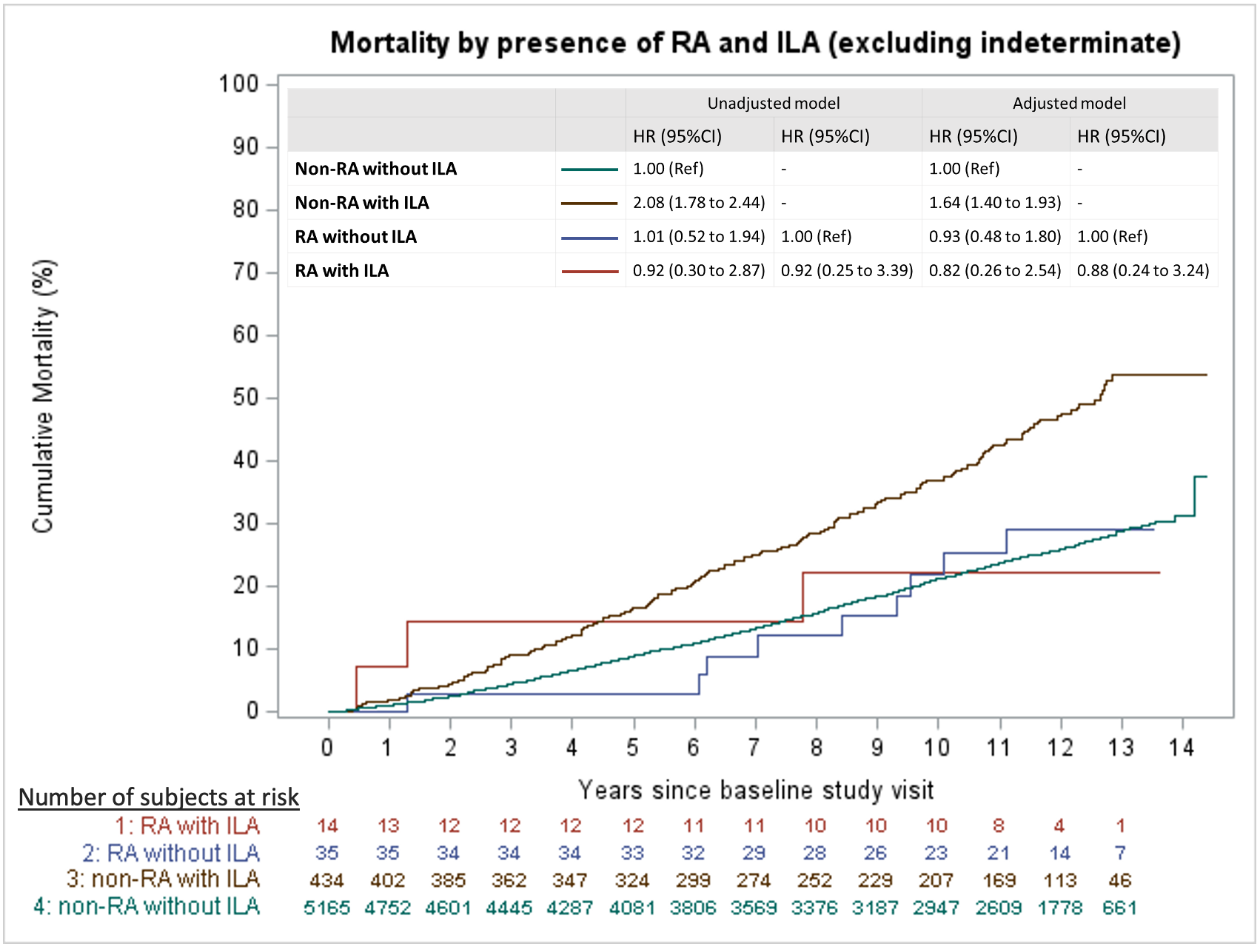

ILA = interstitial lung abnormalities**.** The multivariable model was adjusted for age, sex, smoking status (current/past), pack-years, and body mass index.

**Supplemental Figure S2: Cumulative mortality and hazard ratios stratified by RA/comparator and interstitial lung abnormalities (ILA) status in COPDGene, definite ILA excluded (n=8185)**

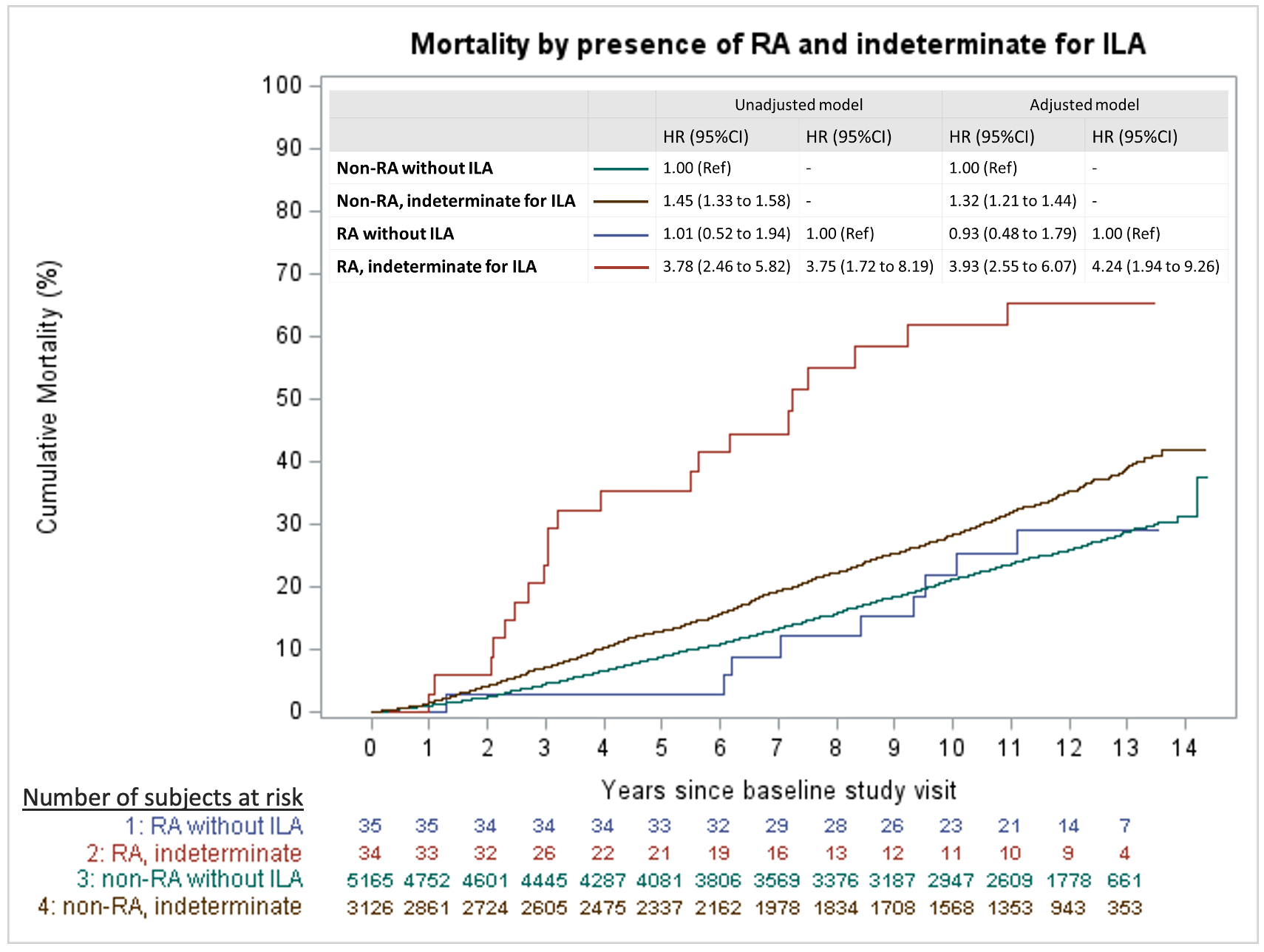
 ILA = interstitial lung abnormalities**.** The multivariable model was adjusted for age, sex, smoking status (current/past), pack-years, and body mass index.
